# Female-male differences in COVID vaccine adverse events have precedence in seasonal flu shots: a potential link to sex-associated baseline gene expression patterns

**DOI:** 10.1101/2021.04.01.21254798

**Authors:** AJ Venkatakrishnan, Praveen Kumar-M, Eli Silvert, Enrique Garcia-Rivera, Mariola Szenk, Rohit Suratekar, Patrick Lenehan, Emily Lindemer, John C. O’Horo, Amy W. Williams, Andrew D. Badley, Abinash Virk, Melanie D. Swift, Gregory J. Gores, Venky Soundararajan

## Abstract

Nearly 150 million doses of FDA-authorized COVID vaccines have been administered in the United States. Sex-based differences of adverse events remain poorly understood, mandating the need for real-world investigation from Electronic Health Records (EHRs) and broader epidemiological data sets. Based on an augmented curation of EHR clinical notes of 31,064 COVID-vaccinated individuals (19,321 females and 11,743 males) in the Mayo Clinic, we find that nausea and vomiting were documented significantly more frequently in females than males after both vaccine doses (nausea: RR_Dose 1_ = 1.67, p_Dose 1_ <0.001, RR_Dose 2_ = 2.2, p_Dose 1_ < 0.001; vomiting: RR_Dose 1_ = 1.58, p_Dose 1_ < 0.001, RR_Dose 2_ = 1.88, p_Dose 1_ = 3.4×10^−2^). Conversely, fever, fatigue, and lymphadenopathy were more common in males after the first dose vaccination (fever RR = 0.62; p = 8.65×10^−3^; fatigue RR = 0.86, p = 2.89×10^−2^; lymphadenopathy RR = 0.61, p = 3.45×10^−3^). Analysis of the Vaccine Adverse Events Reporting System (VAERS) database further confirms that nausea comprises a larger fraction of total reports among females than males (RR: 1.58; p<0.001), while fever comprises a larger fraction of total reports among males than females (RR: 0.84; p<0.001). Importantly, increased reporting of nausea and fever among females and males, respectively, is also observed for prior influenza vaccines in the VAERS database, establishing that these differences are not unique to the recently developed COVID-19 vaccines. Investigating the mechanistic basis underlying these clinical findings, an analysis of bulk RNA-sequencing data from 12,158 human blood samples (8626 female, 3532 male) reveals 85 genes that are not only significantly different in their gene expression between females and males at baseline, but also have established literature-based associations to COVID-19 as well as the vaccine-related adverse events of clinical consequence. The NLRP3 inflammasome and the NR3C1 glucocorticoid receptor emerge as particularly promising baseline links to sex-associated vaccine adverse events, warranting targeted investigation of these signaling pathways and associated cell types. From a public health standpoint, our clinical findings shall aid in educating patients on the sex-associated risks they should expect for COVID-19 vaccines and also promote better clinical management of vaccine-associated adverse events.

## Introduction

Different types of COVID vaccines are being authorized across the world and mass vaccination efforts are underway. Recently, we and others analyzed the safety^1^ and effectiveness^2^ of the Pfizer/BioNtech and the Moderna COVID-19 vaccines based on the analysis of patient records from large health systems. Although males and females were similarly represented in the safety cohorts of the COVID-19 vaccine trials, the safety and tolerability outcomes were only reported at the whole cohort level without a discussion of differences between sexes. Monitoring conducted as part of the U.S. vaccination program reported that the most reported adverse events involving persons not residing in long-term care facilities were women^3^. However, whether there are any differences in the adverse events between men and women remains unclear, warranting a real-world data-based analysis of the adverse event profiles.

It is well-known that vaccination outcomes differ by sex which may be explained by various genetic, immunologic, and hormonal distinctions^4^. In a recent literature review adverse events to the influenza vaccine were reported to be higher in females over males^5^. The availability of vaccine associated adverse events from Mayo Clinic’s multi-state healthcare system provides an excellent opportunity to systematically investigate where there are any sex-specific differences in the COVID vaccine associate adverse events. We have previously developed augmented curation methods that facilitate rapid and real-time extraction of phenotypic data from the Mayo Clinic^6^. In addition, the vaccine adverse events reported in the Vaccine Adverse Event Reporting System (VAERS) is also a larger and diverse resource complimentary to the data present in patient records. Here, we leverage the augmented curation methods on Mayo Clinic patient records along with the analysis of the VAERS database to assess the sex-based differences in the COVID-19 vaccine associated adverse event profiles.

## Methods

### Study Population

The study involves assessment of two sets of data: (i) the individuals from Mayo Clinic Electronic Health Records (EHR) and (ii) the analysis of the adverse events reports submitted in the VAERS online database.

For the dataset from Mayo Clinic Health systems, we identified individuals who underwent polymerase chain reaction (PCR) testing at the Mayo Clinic and hospitals affiliated with the Mayo Clinic Health System. This study was reviewed by the Mayo Clinic Institutional Review Board (IRB) and determined to be exempt from the requirement for IRB approval (45 CFR 46.104d, category 4). Subjects were excluded if they did not have a research authorization on file. The cohorts of vaccinated inclusion in this study are identical to the cohorts considered in our previous analysis on the safety^7^ and effectiveness^2^ of vaccines. The following inclusion criteria were used in these studies: (1) at least 18 years old; (2) no positive SARS-CoV-2 PCR test before December 1, 2020; (3) resides in a locale (based on Zip code) with at least 25 individuals who have received BNT162b2 or mRNA-1273. This data set did not include Ad26.COV2.S (Johnson and Johnson vaccine). Individuals with zero follow-up days after vaccination (i.e. those who received the first vaccine dose on the date of data collection) were also excluded, leaving 31,064 individuals in the final vaccinated cohort. Overall, we mainly assessed a cohort of 31,064 individuals who were administered with a COVID-19 vaccine at the Mayo Clinic Health system between the time period December 15, 2020 and February 8, 2021. 31,064 individuals (19,321 females, 11,743 males) had received the first dose of the vaccine and 17,063 individuals had received the second dose of the vaccine.

#### Definition of adverse effects of interest

We followed the adverse effects described in our prior study^7^, which were primarily derived from those assessed in Phase III trials of BNT162b2 and mRNA-1273^8,9^, including fatigue, fever, chills, myalgia, arthralgia, headache, lymphadenopathy, erythema, diarrhea, vomiting, and local pain and swelling. Anaphylaxis and facial paralysis (Bell’s palsy) were also included as these rare events have been reported in individuals receiving COVID-19 vaccines as well^10,11^. We mapped each adverse effect to a set of synonyms intended to capture the most common ways that a given phenotype would be referenced in the context of a clinical note.

#### Curation of adverse effects from clinical notes

To curate the adverse effects experienced by each patient from the electronic health record, we used a Bidirectional Encoder Representations from Transformers (BERT)-based neural network model^12^ to classify the sentiment for the phenotypes (described above) mentioned in the clinical notes. Specifically, this classification model categorizes phenotype-containing sentences into one of four categories: (1) confirmed diagnosis, (2) ruled-out diagnosis, (3) possibility of disease, and (4) alternate context (e.g. family history). This classification model was trained on 18,500 sentences and has shown an out-of-sample accuracy of 93.6% with precision and recall scores above 95%^6^. For each individual, we applied the sentiment model to the clinical notes in the Mayo Clinic electronic health record during our defined intervals of interest: (1) Day V_1_ to 21 days after Day V_1_, and (3) Day V_2_ to 21 days after Day V_2_. For each phenotype, we identified the first date on which the given individual had at least one sentence in which the phenotype was categorized as “confirmed diagnosis” with a confidence score of at least 90%. For the severe phenotype anaphylaxis, each such sentence was manually reviewed to verify the positive sentiment (i.e. confirmed diagnosis) and to assess the tense of this sentiment (i.e. past vs. present). Only sentences which confirm a present diagnosis were used to count anaphylaxis events in this study.

#### Curation of adverse effects from VAERS database

We obtained the adverse events related to COVID-19 vaccine for all three approved vaccines in the US (Pfizer/BioNtech - BNT162b2, Moderna - mRNA-1273 and Janssen - Ad26.CoV2.S) from the VAERS database. Our inclusion criteria for considering an adverse event for analysis was i) the onset of symptom date for the adverse event should be between December 15th 2020 to March 12th 2021. We removed (i) mislabeled adverse events where symptoms occurred earlier than December 15, 2020, ii) reports which were filed without a specified vaccine manufacturer, iii) adverse events whose date of onset of symptom was before the date of vaccination iv) adverse events which were filed without a specified gender, and v) adverse events which were obviously not untoward/abnormal. The frequencies of the adverse events were computed at the level of unique VAERS id. We considered all the adverse events which fit under these criteria but we restricted our focus only to the adverse events phenotype described above. For the comparator, we also obtained the adverse events for influenza (seasonal) flu vaccine. The following Flu vaccine types were considered: ‘FLUR3’, ‘FLUN3’, ‘FLU4’, ‘FLUA3’, ‘H5N1’,’FLU3’, ‘FLUC4’, ‘FLUX’, ‘FLUR4’, ‘FLUN4’, ‘FLUC3’. We assessed the data separately for 2019 (January 1st 2019 to December 31st 2019), 2020 (January 1st 2020 to January 31th 2020). We applied the inclusion criteria iii, iv and v described above for COVID-19 vaccine to the flu vaccines and filtered out the adverse events. The rest of the analysis for the Flu vaccine are the same as that of COVID-19 vaccine. We stratified the adverse events profiles based on gender and age (<55 years and ≥ 55 years).

#### Statistical analysis

To assess the probability of an adverse event for a given demographic, we calculated relative risk (RR) ratios for patients experiencing adverse events reported across cohorts, grouped by sex.

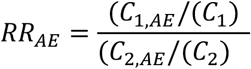

Where,

RR_AE_ = Risk Ratio of Adverse Event appearing in Group 1 over Group 2

C_1AE_ = Number of Adverse Event for Group 1

C_1_ = Total Group 1 Population

C_2AE_ = Number of Adverse Events for Group 2

C_2_ = Total Group 2 Population

We calculated 95% adjusted confidence intervals for relative risk ratios by the adjusted Wald method.

To test significance between groups, we calculated Benjamini-Hochberg adjusted χ2 p-values for adverse events. The contingency table constructed for the analysis was

|  | Specific AE (e.g. nausea) | Without AE |
| --- | --- | --- |
| **Females** | Number of vaccinated female patients with the AE (e.g. nausea) | Number of vaccinated female patients without the AE (e.g. nausea) |
| **Males** | Number of vaccinated male patients with the AE | Number of vaccinated males without the AE |

For comparison of the proportion of vaccine-related adverse events in Mayo Clinic with VAERS, we computed a *conditional RR* (cRR) with the denominator as the number of vaccinated females or vaccinated males reporting any adverse event.

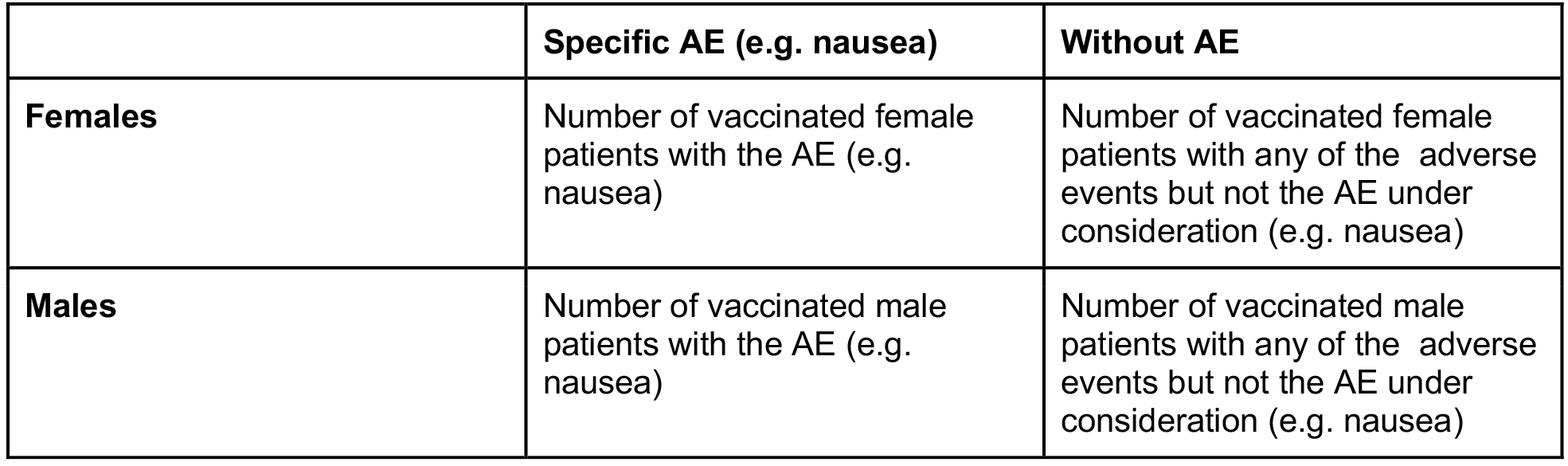

#### Comparison of expression levels of all genes at baseline in blood samples of males and females

Gene expression data was processed from raw sequencing files obtained from the Gene Expression Omnibus (GEO)^13^. Briefly, our pipelines consisted of pre-processing FASTQ files with *fastp*^*14*^ and gene expression quantification using GRCh38 human genome as reference with *salmon*^*15*^. For quantification we utilized Transcripts per Million (TPM) as the gene expression values. Metadata was scraped from the GEO database and entity type was extracted from the textual characteristics field using an internal Named Entity Recognition service. This provided labels for both Blood-related and sex-related categories. All downstream analyses were performed in R. Significance for gene expression difference were calculated using Welch’s t-test, and p-values were corrected using the Bonferroni method. Literature-based associations were calculated using conditional probabilities of co-occurrence in the biomedical literature (incl. PubMed abstracts, PMC full articles, pre-prints, grants, patents and media articles), and have been described in detail elsewhere^16^.

## Results

To identify COVID-19 vaccine associated adverse events which may be disproportionately experienced between males and females, we performed augmented curation of EHR notes from 19321 females and 11743 males who received at least one dose of either BNT162b2 or mRNA-1273 at the Mayo clinic. Of these, 1735 females and 1052 males reported at least one adverse event within 21 days following the first vaccine dose. Among 11483 females and 5580 males who received two doses of either vaccine, 727 females and 294 males reported at least one event within 21 days after the second dose. Nausea and vomiting were more documented more frequently in females than males after both doses (nausea: RR_Dose 1_ = 1.67, p_Dose 1_ = 2.90×10^−9^, RR_Dose 2_ = 2.2, p_Dose 1_ = 2.68×10^−6^; vomiting: RR_Dose 1_ = 1.58, p_Dose 1_ = 7.95×10^−4^, RR_Dose 2_ = 1.88, p_Dose 1_ = 0.034; **Table 1**), while fever, fatigue, and lymphadenopathy were more common in males (fever RR = 0.62; p = 8.65×10^−3^; fatigue RR = 0.86, p = 2.89×10^−2^; lymphadenopathy RR = 0.61, p = 3.45×10^−3^; **Table 1**). Other adverse events were experienced at similar rates among females and males, including arthralgia, myalgia, and headache (**Table 1**).

**Table 1:** Sex-based risk ratios of experiencing AEs after 1st and 2nd shots (based on symptoms extracted from Mayo Clinic EHRs).

| AE | % of females who experienced AE after 1st shot out of all females who got 1st shot (n = 19321) | % of males who experienced AE after 1st shot out of all males who got 1st shot (n = 11743) | F/M risk ratio (exact 95% CI) (after 1st shot) | Corrected P-value | % of females who experienced AE after 2nd shot out of all females who got 2nd shot (n = 11483) | % of males who experienced AE after 2nd shot out of all males who got 2nd shot (n = 5580) | F/M risk ratio (exact 95% CI) (after 2nd shot) | Corrected P-value |
| --- | --- | --- | --- | --- | --- | --- | --- | --- |
| Nausea | 2.85% (n = 550) | 1.7% (n = 200) | 1.67 (1.42, 1.98) | 2.90E-09 | 1.97% (n = 226) | 0.9% (n = 50) | 2.2 (1.61, 3.05) | 2.68E-06 |
| Vomiting | 1.31% (n = 254) | 0.83% (n = 98) | 1.58 (1.24, 2.01) | 7.95E-04 | 0.81% (n = 93) | 0.43% (n = 24) | 1.88 (1.19, 3.09) | 3.36E-02 |
| Soreness | 0.67% (n = 130) | 0.43% (n = 50) | 1.58 (1.13, 2.24) | 1.62E-02 | 0.53% (n = 61) | 0.34% (n = 19) | 1.56 (0.92, 2.77) | 3.74E-01 |
| Local pain | 0.04% (n = 7) | 0.03% (n = 3) | 1.42 (0.32, 8.5) | 7.34E-01 | 0.01% (n = 1) | 0.04% (n = 2) | 0.24 (0.0, 4.67) | 4.21E-01 |
| Myalgia | 1.05% (n = 203) | 0.89% (n = 104) | 1.19 (0.93, 1.52) | 3.30E-01 | 0.72% (n = 83) | 0.57% (n = 32) | 1.26 (0.83, 1.96) | 4.21E-01 |
| Diarrhea | 1.49% (n = 287) | 1.37% (n = 161) | 1.08 (0.89, 1.32) | 7.33E-01 | 0.65% (n = 75) | 0.86% (n = 48) | 0.76 (0.52, 1.11) | 3.74E-01 |
| Erythema | 1.34% (n = 259) | 1.28% (n = 150) | 1.05 (0.85, 1.29) | 7.34E-01 | 0.82% (n = 94) | 0.81% (n = 45) | 1.02 (0.7, 1.48) | 9.34E-01 |
| Arthralgia | 1.46% (n = 282) | 1.48% (n = 174) | 0.99 (0.81, 1.2) | 8.75E-01 | 0.98% (n = 113) | 0.9% (n = 50) | 1.1 (0.78, 1.56) | 8.11E-01 |
| Headache | 0.06% (n = 12) | 0.07% (n = 8) | 0.91 (0.34, 2.57) | 8.75E-01 | 0.09% (n = 10) | 0.04% (n = 2) | 2.43 (0.52, 22.81) | 4.21E-01 |
| Chills | 0.39% (n = 75) | 0.43% (n = 51) | 0.89 (0.62, 1.3) | 7.33E-01 | 0.36% (n = 41) | 0.32% (n = 18) | 1.11 (0.62, 2.05) | 8.32E-01 |
| Fatigue | 3.54% (n = 683) | 4.1% (n = 481) | 0.86 (0.77, 0.97) | 2.89E-02 | 2.3% (n = 264) | 1.92% (n = 107) | 1.2 (0.95, 1.52) | 3.74E-01 |
| Facial paralysis | 0.04% (n = 7) | 0.05% (n = 6) | 0.71 (0.2, 2.55) | 7.33E-01 | 0.03% (n = 3) | 0.02% (n = 1) | 1.46 (0.12, 76.53) | 8.32E-01 |
| Fever | 0.42% (n = 82) | 0.68% (n = 80) | 0.62 (0.45, 0.86) | 8.65E-03 | 0.26% (n = 30) | 0.36% (n = 20) | 0.73 (0.4, 1.35) | 4.21E-01 |
| Lymphadenopathy | 0.49% (n = 95) | 0.8% (n = 94) | 0.61 (0.46, 0.83) | 3.45E-03 | 0.33% (n = 38) | 0.36% (n = 20) | 0.92 (0.52, 1.67) | 8.32E-01 |
| Local swelling | 0.02% (n = 3) | 0.03% (n = 3) | 0.61 (0.08, 4.54) | 7.33E-01 | 0.0% (n = 0) | 0.0% (n = 0) | inf (nan, inf) | nan |
| Anaphylaxis | 0.01% (n = 1) | 0.0% (n = 0) | 0.0 (nan, inf) | nan | 0.0% (n = 0) | 0.0% (n = 0) | inf (nan, inf) | nan |

**Table 2.**
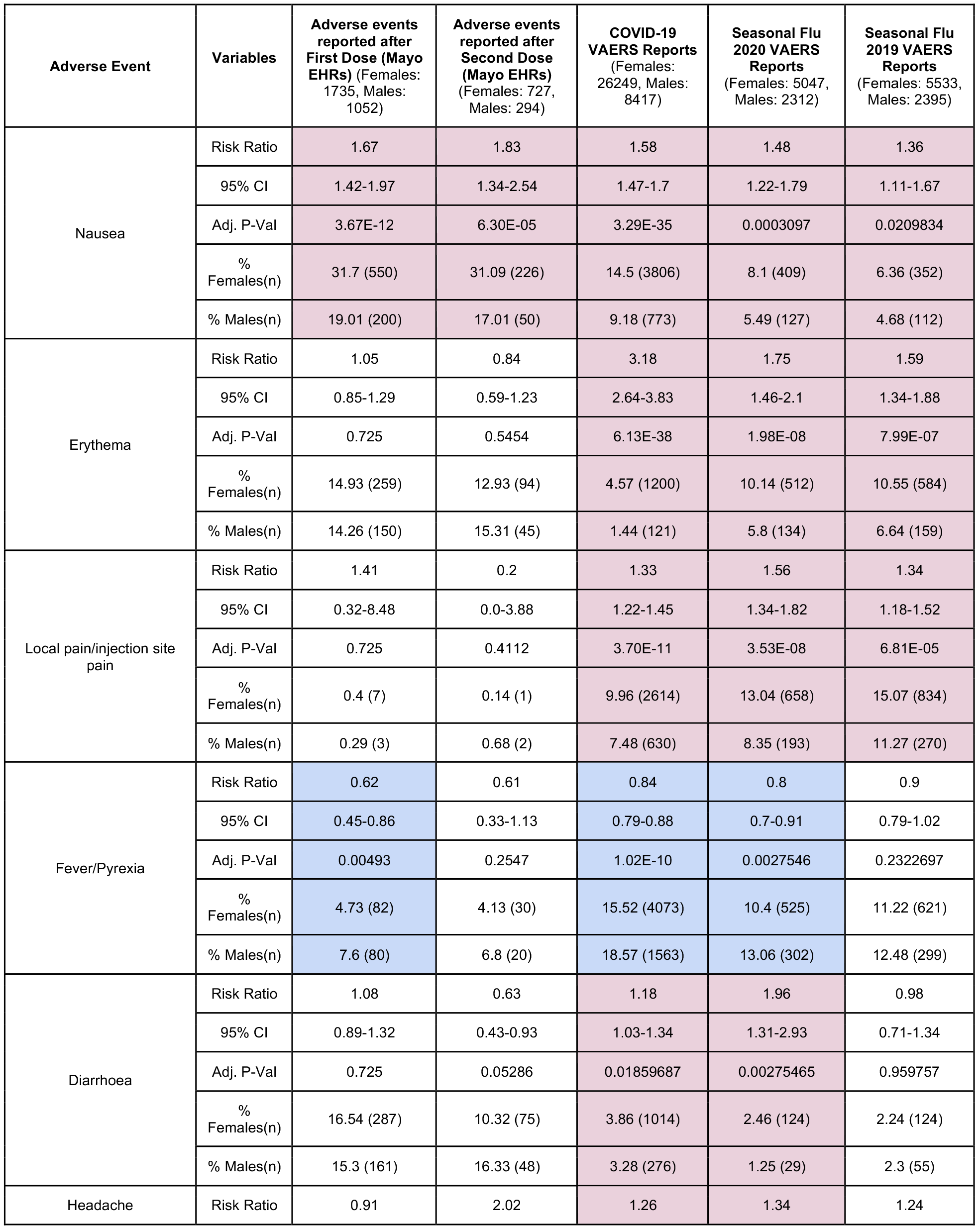
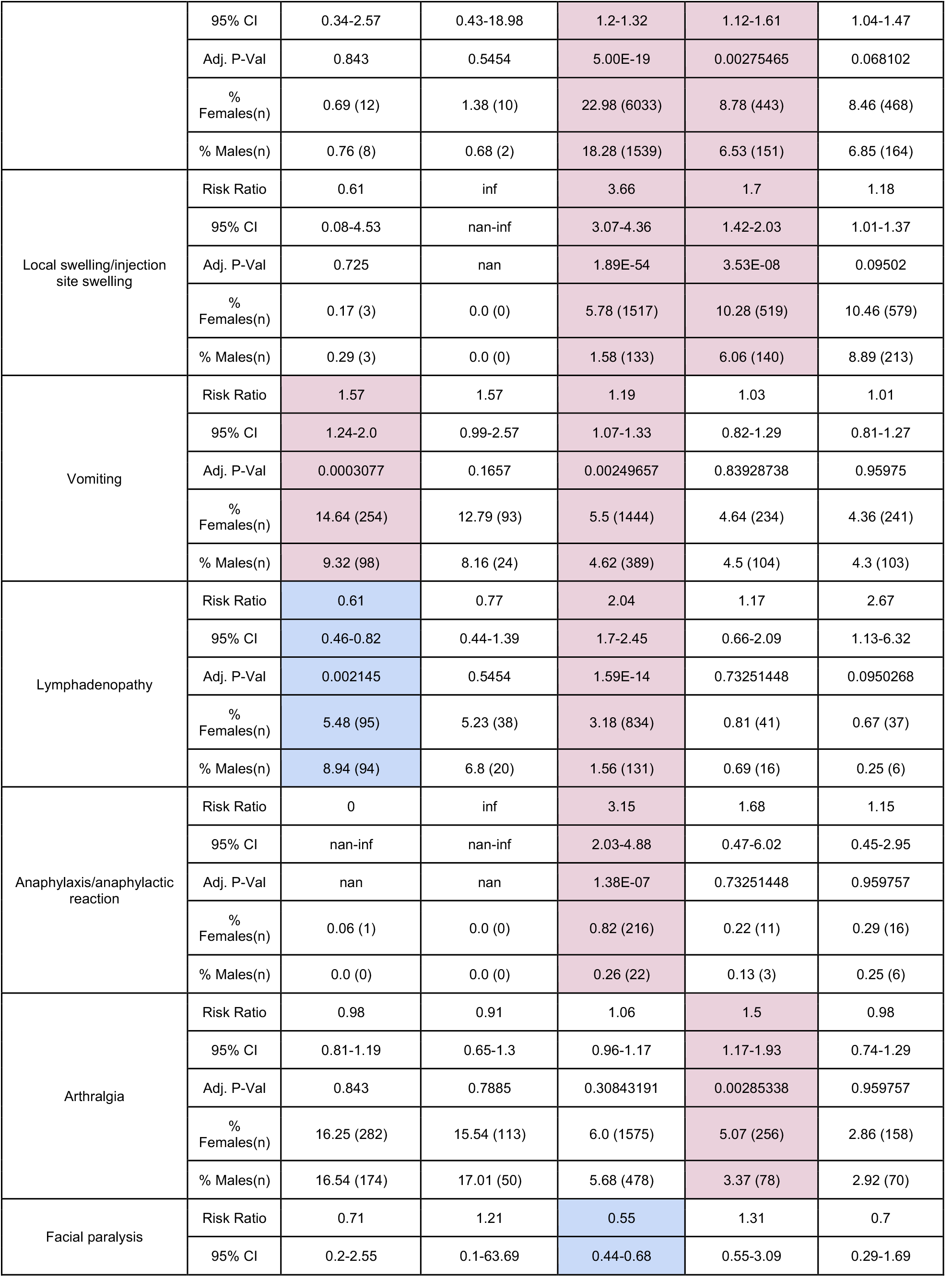
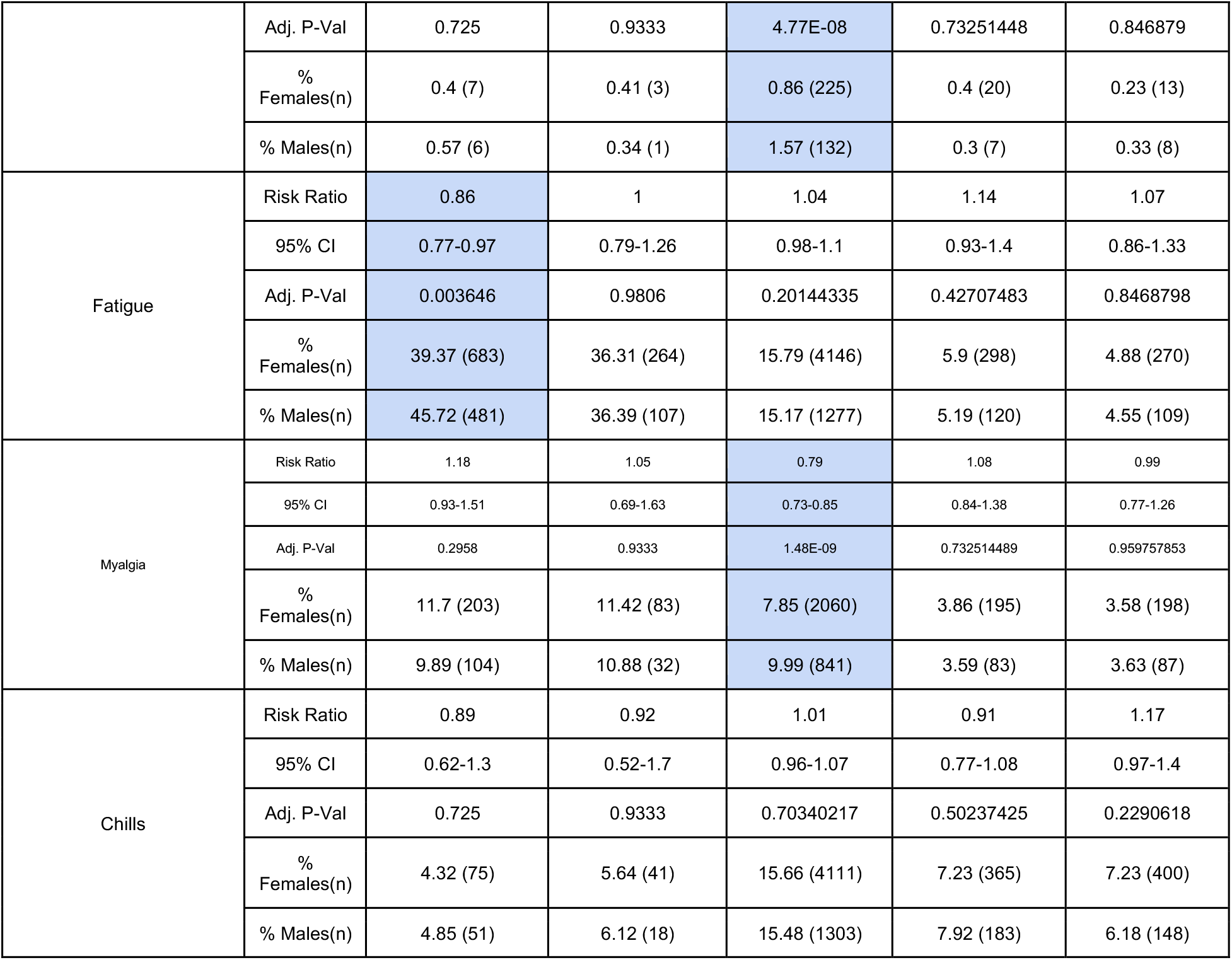
Sex-based risk ratio of reporting AEs across the Mayo Clinic and VAERS. Purple-colored rows are biased towards females (Risk Ratio > 1 AND adjusted P-Value < 0.05) and blue-colored rows are biased towards males (Risk Ratio < 1 AND adjusted P-Value < 0.05).

One limitation of studying vaccine associated adverse events through EHR curation is that it will only capture events in individuals who experience side effects serious enough to warrant clinical attention or who happen to have a routine clinical visit shortly after their vaccination. Alternatively, any vaccinated individual can report their experience of adverse events through the FDA Vaccine Adverse Events Reporting System (VAERS), even outside of the clinical care context. To triangulate our observations from the Mayo Clinic system, we thus analyzed the 34,666 COVID-19 vaccine associated adverse event reports (26,249 females and 8,417 males) in the VAERS database. Since the data we analyzed from VAERS is based on reports that had any adverse event, we compared the adverse events in females and males from Mayo Clinic as a proportion of vaccinated females and males that reported any adverse event (conditional RR; cRR). Consistent with our EHR-based analysis, reports of nausea and vomiting comprised a larger fraction of reports from females than males (nausea cRR: 1.58, p<0.001; vomiting cRR: 1.19, p = 0.002; **Table 1**). Conversely, reports of fever/pyrexia comprised a larger fraction of reports from males than females (cRR = 0.84; p<0.001). Several other adverse events were disproportionately reported in females (e.g. erythema, pain, diarrhea, headache, anaphylaxis) or males (e.g. facial paralysis, myalgia) in VAERS but not per EHR curation. These discrepancies may be due to differences in the underlying populations and sources of bias in the reporting and recording of adverse events through these systems.

To understand whether the predisposition of females to experience nausea and vomiting, or predisposition of males to experience fever, after receiving the COVID-19 vaccine should be viewed as surprising or concerning, we performed a similar analysis of VAERS reports associated with the influenza vaccine in 2019 and 2020. Interestingly, nausea comprised a higher fraction of these reports among females than males in both years, and fever comprised a higher fraction of these reports among males in 2019. Several other effects which were more prevalent among COVID-19 vaccine associated reports from females (e.g. erythema, local pain, diarrhea, and headache) were also disproportionately reported among females after receiving the flu vaccine in 2020 and/or 2019.

Taken together, this triangulation effort highlights that while there are indeed likely sex-based differences in the adverse event profiles associated with COVID-19 vaccination. Specifically, our analyses of a single EHR and a national surveillance database highlight nausea and fever as side effects that disproportionately affect females and males, respectively. It is important to note, however, that these differences mirror prior experience with flu vaccines and thus should not be portrayed as phenomena which are specific to or particularly concerning for the recently developed COVID-19 vaccines.

Finally, in order to identify potential mechanisms behind sex-associated sensitivity to the COVID vaccines we compared the expression levels of all genes in blood samples of males and females. This data was assembled from a host of studies taken from the Gene Expression Omnibus (GEO) where the sample source was described as blood, and it includes 3,532 male blood samples and 8,626 female blood samples. By comparing these cohorts, we identified 8,257 genes out of ∼60,000 gene-entities from Ensembl that were significantly different (Bonferroni adjusted p-value < 0.05 and log2 Fold Change > 0.5) between males and females (**Supplementary Figure 1**). We next obtained the amount of literature associated with these differential genes to COVID19 and the adverse events that have been reported for the SARS-CoV-2 vaccine. Out of these, 85 genes also had a significant literature-based association to COVID19 and the vaccine related adverse events. Several of these genes stand out as potential mechanisms for sex-specific sensitivity to the vaccine. Interestingly, we found the inflammasome (through NLRP3 expression) to be higher in males compared to females and in the top differential genes (**Supplementary Figure 2**) and has a strong association with ‘fever’ in literature. This is concordant with the observation from the analysis of adverse events from Mayo Clinic data and VAERS, motivating the need to explore whether inflammasome could have a sex-specific role in the adverse events. In the hormone receptor family, we identified the glucocorticoid receptor NR3C1 as being highly expressed in females over males and also having a strong literature association with COVID19 (**Supplementary Figure 3**), fatigue, nausea and vomiting. This gene has been previously implicated in COVID19 severity^17^ and could be linked to the increased nausea and vomiting to the vaccine in females.

## Discussion

Our findings in the current study demonstrate higher reported nausea and vomiting in women, and higher reported fever in men who experience adverse events (AE) related to the COVID-19 vaccine across all datasets assessed. We additionally find that erythema, local pain and swelling, headache, diarrhea, and anaphylaxis have an increased prevalence in females with AEs and facial paralysis and myalgia are increased in men with AEs in the VAERS database. It should be noted that our findings do not demonstrate that women are more likely to experience any adverse events, as we did not assess the entire population who received the COVID vaccine. Rather, our findings suggest that there is a wider range of AEs that women are more likely to experience than men, if they experience any at all. While these findings are important for public health messaging, it is also important to note that they are similar to findings seen before with the influenza vaccine, and therefore should not cause reason for vaccine fear or skepticism.

There are several potential explanations for the differences in the adverse event profiles of females and males. A recent review of vaccine-induced hormonal immunity suggests that women experience enhanced immune reactogenicity, leading them to be more immune to infectious diseases, but also resulting in a higher rate of adverse events^18^. These sex-based differences could be do to hormonal, genetic, and microbiota responses, with a particular emphasis on the role of sex hormones and immune response to vaccines. Indeed, interactions between estrogen and flu vaccines have been hypothesized to boost immunity in other studies^19,20^, and increased levels of estrogen are linked to both nausea and vomiting^21^, the two most salient adverse events that we see in the female population.

In addition to biological reasons for sex-based differences in AE profiles, there are explanations for these differences that are related differences between men and women in the way that they perceive pain and how they interact with the healthcare system^22^. There is a growing body of literature to suggest that men and women experience pain and noxious stimuli differently, with women showing a greater sensitivity to pain.^23–25^ This is in line with our findings, where the more female-dominant AEs are those related to pain (headache, local soreness, and even nausea) and the more male-dominant AEs are not associated with pain. A more behavioral explanation for our findings relates to sex-based differences in utilization of the healthcare system. It is well-known that men and women are treated differently by the healthcare system, and this in turn leads to sex-based differences in overall utilization. Women tend to use outpatient care and self-report lower health statuses than men^2627^, and while men tend to suffer more from severe and chronic conditions, they are much less likely to seek treatment for or report acute symptoms^28^. Our findings are likely due to a combination of all of these factors.

There could be potential confounders for the sex-associated adverse events observed in this study. These include pre-existing conditions, prior history of COVID, demographic characteristics such race/ethnicity and vaccination history of other vaccines. Future studies accounting for these demographic variables and patient history would help delineate the sex-associated differences in adverse events. The dataset from Mayo Clinic was limited to individuals who had undergone PCR testing for SARS-CoV-2, and this could introduce potential confounders related to the reason for testing, such as an illness, planned procedure, or travel.

Overall, though there are differences in the COVID vaccine associated adverse event profiles between females and males, such differences have been seen in Flu vaccines previously. Our findings will help in educating the patients and healthcare practitioners on the expected sex-specific risks to COVID vaccines as well as managing these adverse events prophylactically or therapeutically. With the roll-out of new COVID vaccines and the observation of sex-associated side effects such as cerebral venous sinus thrombosis (CVST) seen in some younger women after receiving the AstraZeneca vaccine^29^, there is a need to monitor the adverse events for all the COVID vaccines in near real-time in order to guide public health policies. Finally, the findings here motivate the need for clinical studies on causality and the biological mechanisms underlying the differences in the sex-associated adverse event profiles for COVID-19 vaccines.

## Data Availability

After publication, the data will be made available upon reasonable requests to the corresponding author. A proposal with detailed description of study objectives and the statistical analysis plan will be needed for evaluation of the reasonability of requests. Deidentified data will be provided after approval from the corresponding author and the Mayo Clinic.

## Author Contributions

VS and AJV conceived the study. AJV, PL, MS, EL, PK and VS wrote the manuscript and reviewed the findings. AJV, PK, ES, MS, RS, PL and EG, contributed methods, analysis, and software. JCOH, GJG, AWW, ADB, MDS, AV, and JH reviewed the study, findings, and the manuscript. All authors revised the manuscript.

---

**Figure 1.**
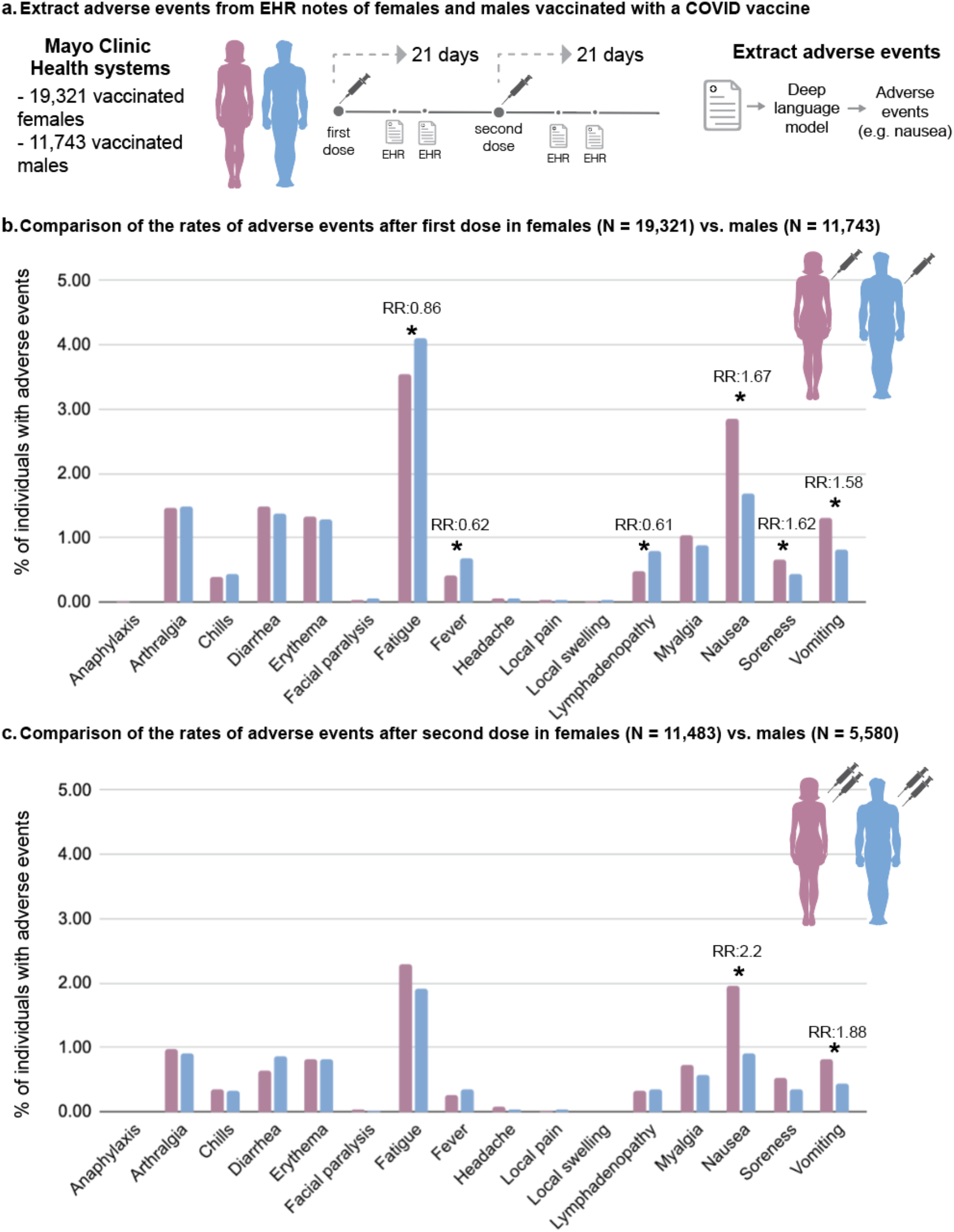
Overview of the comparison of COVID-associated adverse events between females and males extracted from Mayo Clinic EHRs. a. Extract adverse events from females and males vaccinated with a COVID vaccine. b. Comparison of the rates of adverse events after first dose in females (N = 19,321) vs. males (N = 11,743). c. Comparison of the rates of adverse events after the second dose in females (N = 11,483) vs. males (N = 5,580).

## Supplementary Figures

**Supplementary Figure 1.**
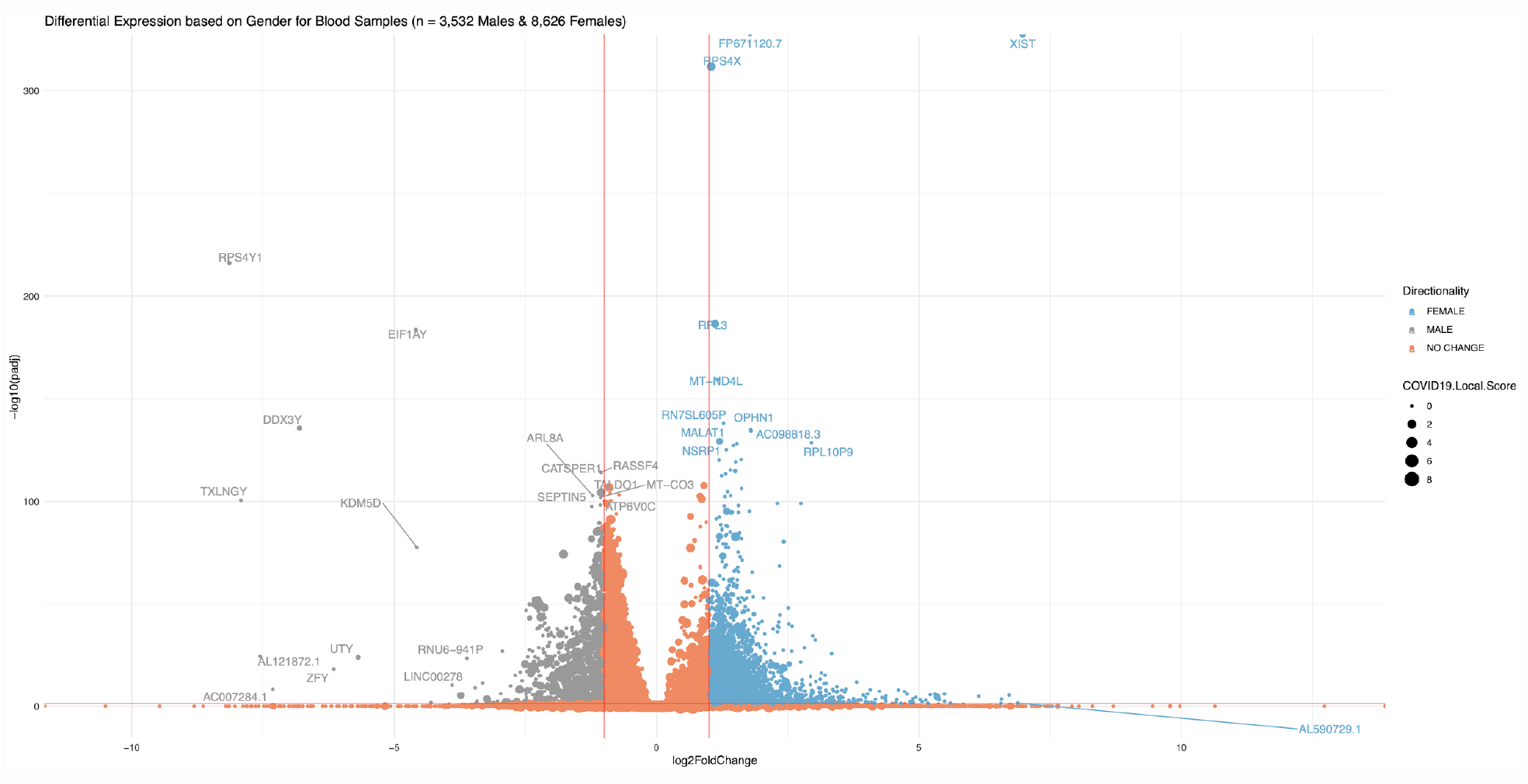
Volcano plot of the differential expression of genes in female vs male blood samples. The x-axis shows the log2 fold change between females and males for the mean Transcript per Million (TPM) expression. Y-axis represents the bonferroni adjusted p-value of a Welch’s t-test between the female and male mean TPM expression. The size of each dot represents the literature association strength between the gene and “COVID19”. Red lines shown at log2 fold change of 1 and adjusted p-value of 0.05.

**Supplementary Figure 2.**
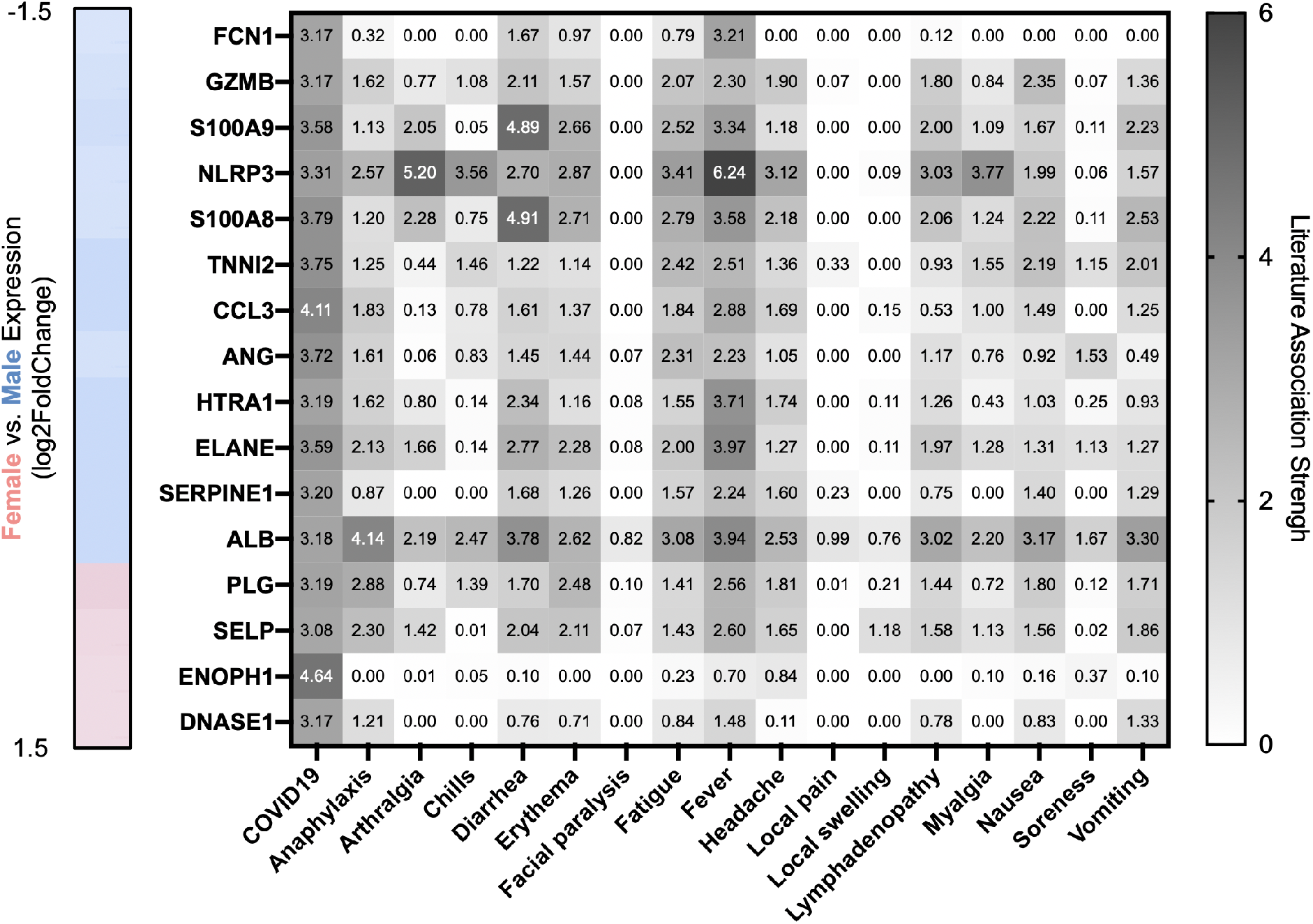
Literature associations between top differential genes (log2 fold change > 1) and the adverse events reported for the SARS-CoV-2 vaccine. Each cell within the heatmap contains the Literature Local Score between the column’s concept and the gene. Also shown on the left of the heatmap is the log2FoldChange for female vs. male expression as a color scale.

**Supplementary Figure 3.**
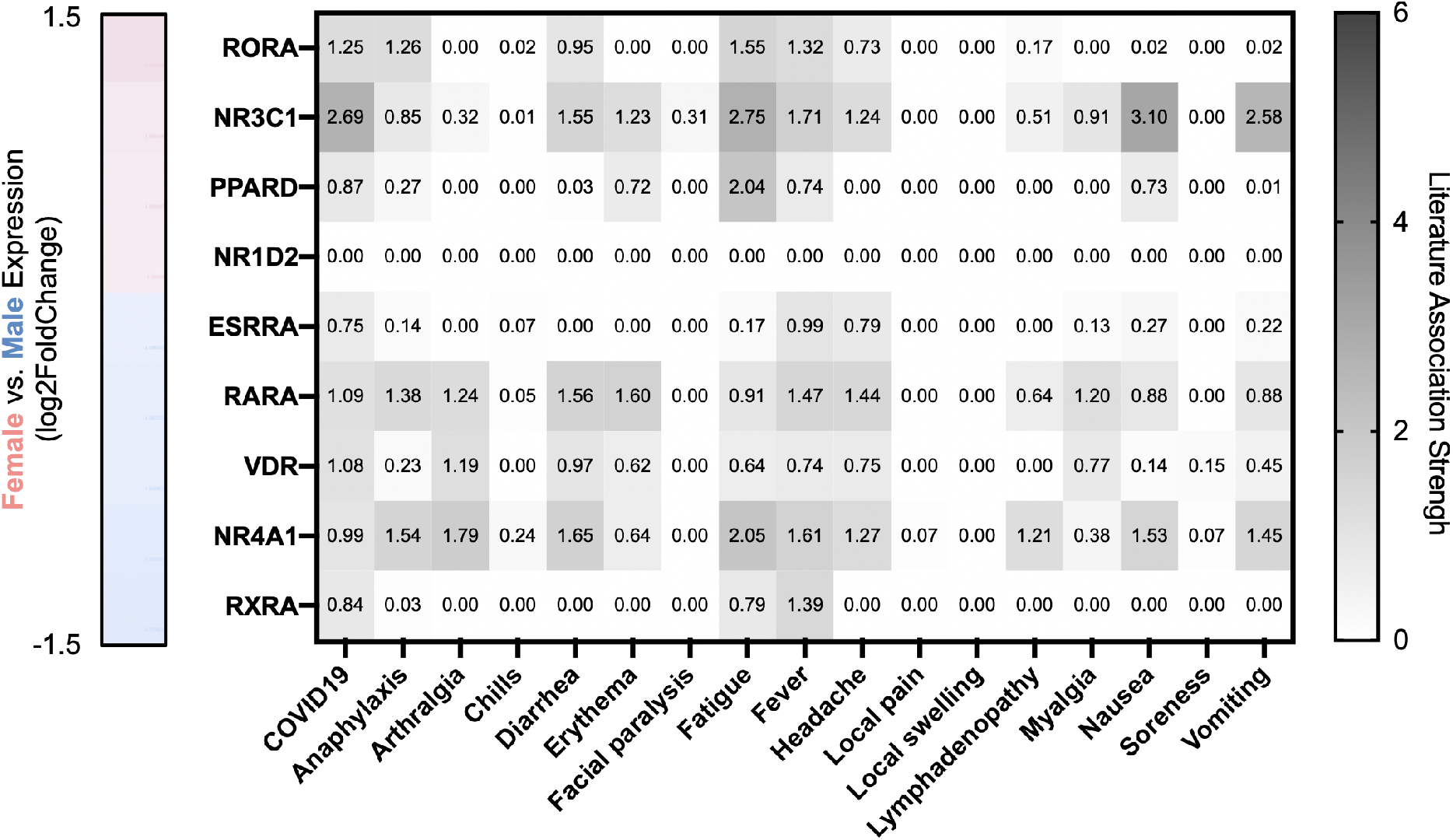
Literature-based associations between hormone receptor genes and the adverse events reported for the COVID vaccines. Each cell within the heatmap contains the Literature Local Score between the column’s concept and the gene. Also shown on the left of the heatmap is the log2FoldChange for female vs. male expression as a color scale.

